# Novel scotoma detection method using time required for fixation to the random targets

**DOI:** 10.1101/2021.06.03.21258101

**Authors:** Nobuyuki Takahashi, Shozo Saeki, Minoru Kawahara, Hirohisa Aman, Eri Nakano, Yuki Mori, Masahiro Miyake, Hiroshi Tamura, Akitaka Tsujikawa

**Affiliations:** Matsuyama School for the Blind, Matsuyama, Ehime, Japan; Center for Information Technology, Ehime University, Matsuyama, Ehime, Japan; Graduate School of Science and Engineering, Ehime University, Matsuyama, Ehime, Japan; Business Strategy Dept, FINDEX Inc., Matsuyama, Ehime, Japan; Department of Ophthalmology and Visual Sciences, Kyoto University Graduate School of Medicine, Kyoto, Japan; Center for Innovative Research and Education in Data Science, Institute for Liberal Arts and Sciences, Kyoto University

## Abstract

We developed a novel scotoma detection system using time required for fixation to the random targets, or the” eye-guided scotoma detection method “. In order to verify the” eye-guided scotoma detection method “, we measured 78 eyes of 40 subjects, and examined the measurement results in comparison with the results of measurement by Humphrey perimetry. The results were as follows: (1) Mariotte scotomas were detected in 100% of the eyes tested; (2) The false-negative rate (the percentage of cases where a scotoma was evaluated as a non-scotoma) was less than 10%; (3) The positive point distribution in the low-sensitivity eyes was well matched. These findings suggested that the novel scotoma detection method in the current study will pave the way for the realization of mass screening to detect pathological scotoma earlier.

**Author summary:** Conventional perimeters, such as the Goldmann perimeter and Humphrey perimeter, require experienced examiners and space occupying. With either perimeter, subjects’ eye movements need to be strictly fixed to the fixation target of the device. Other perimeters can monitor fixation and automatically measure the visual field. With the eye-guided scotoma detection method proposed in the current study, subjects feel less burdened since they do not have to fixate on the fixation target of the device and can move their eyes freely. Subjects simply respond to visual targets on the display; then, scotomas can be automatically detected. The novel method yields highly accurate scotoma detection through an algorithm that separates scotomas from non-scotomas.

## Introduction

### The relationship between visual field loss and quality of life

Various diseases, including glaucoma, cause pathological scotoma. However, scotoma is often difficult to detect, and it might advance to an untreatable stage if detected too late. Like visual acuity, visual field loss is an important factor in visual function assessment. Moreover, visual field loss is deeply related to quality of life (QoL) [1–3]. According to an estimate by the World Health Organization, at least 2.2 billion people globally have a vision impairment, including 1 billion people whose impairment has yet to be addressed [4]. Globally, major causative diseases of blindness among adults aged 50 years or older are cataract, uncorrected refractive error, and glaucoma [5]. The impact of vision impairments, including visual acuity loss and visual field loss, on QoL, is deeply related to the severity, location of the scotoma, and the condition of binocular vision [2, 6, 7]. Hence, early detection and appropriate follow-up of vision impairments are essential to maintain the QoL. However, because the visual field is compensatory (i.e., the other eye compensates with information) and complementary (i.e., the visual center complements a defect with the information around the defect), the presence of an initial visual field loss is not recognized consciously. Therefore, periodic inspection is important for the early detection of visual field loss.

### The necessity of quantitative measurement of the visual field

Quicker and repeatable image technologies for visual field assessment have recently been proposed, such as optical coherence tomography (OCT). The results of OCT do not rely on subjective responses of the patient and detect earlier stages of the disease [8]. On the other hand, the standard automated perimeters, such as the Humphrey field analyzer (HFA), rely on the patient’s subjective responses and have long test times [9]. However, the standard automated perimeter is the most common method in clinical practice and standard for diagnosis and follow-up [10]. In addition, visual field loss is deeply related to vision-related QoL [1, 2]. Hence, the quantitative measurement of retinal sensitivity, which is needed to assess visual field loss, is important for QoL.

If scotoma detection by mass screening becomes available at schools and workplaces, it will be possible to encourage individuals suspected of having visual field loss to see a specialized medical institution at an early stage. By providing a way to confirm visual field loss, mass screening would be beneficial for maintaining QoL. However, despite the high need for mass screening for visual field losses, there is still no appropriate method for this purpose [11]. In this study, therefore, we propose a new method of scotoma detection that would make mass screening feasible.

## Methods

In this study, we present a new method for eye-guided scotoma detection (or vision-guidance perimetry) that does not require the subject to fixate at the center of the visual field, allowing free eye movement, without a large and complicated system.

The eye-guided scotoma detection method is designed to measure the visual field based on the time the subject requires to recognize and respond to the target. In this method, the subject ‘s sightline is guided to follow a target on a PC screen. The procedure is as follows. First, display a target with a notch drawn in Fig 1 at the center of the visual field (this is called the” fixation point “) (A in Fig 1). Generate the target randomly with a notch in the upper/lower and right/left directions. The response is elicited as follows. When the notch direction is entered by moving a cursor or using an input device, such as a joystick, the target disappears and immediately reappears at a different location in the visual field (this is called the” measuring point “) (B in Fig 1). Then the notch direction is entered again using the input device. The time difference between these two inputs (this is called the” response time “) is measured and recorded, and the sequence of actions is repeated for the entire visual field. If the target is displayed at a scotoma in the visual field after the first input (B in Fig 1), the subject loses the target and needs extra time to find it. This method is designed to determine the scotoma location in the visual field by detecting the time delay.

**Fig 1.**
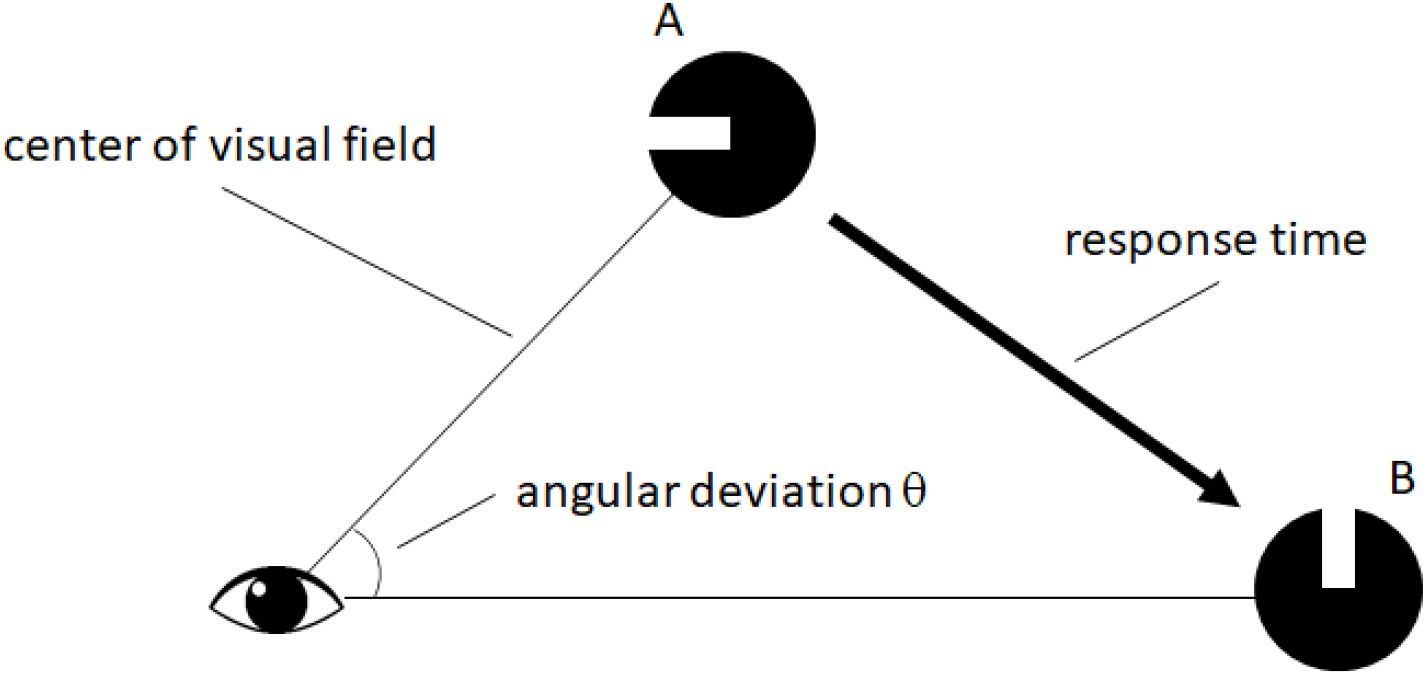
Operation principle of the eye-guided scotoma detection method. Measure the time between the response to the target displayed at the fixation point (A) and the response to the target displayed at the measuring point (B).

For patients with visual field constriction, if the mouse pointer goes out of the visual field during computer operation, they lose the mouse pointer and need time to find it. In such cases, two lines that intersect at the position of the mouse pointer will appear on the screen to guide the patient and reduce the time required to find the mouse pointer. In this situation, the subject also loses the mouse pointer if it is displayed at a scotoma, even in the visual field. Thus, it takes time to capture the mouse pointer in the non-scotoma area by moving the sightline, causing a delay before the next movement starts. We reasoned that the presence of a scotoma could be determined by detecting this delay. We, therefore, decided to replace the mouse pointer with a target to measure the time between the target display in the visual field and the response. When the subject is fixated on the target, the target will be perceived at the center of the visual field. When the target disappears and immediately reappears at a different position, the subject can respond immediately if he/she has no visual field defect at the target position; otherwise, there must be a delay. If this sequence of actions is repeated over the entire visual field, we will identify the scotoma’s position in the visual field. When setting up a gazing point at a position on the display screen that is directly opposite to the subject’s eyes, and displaying the target at the gazing point, some method must be devised to prevent the subject from responding if he/she does not recognize the target. In this article, we propose that, by making a notch on the circular target and instructing the subject to enter the notch direction, the sightline can be guided naturally to the fixation point because the subject must look at the visual field center to recognize the notch. At the position of scotoma detection (measuring point), it is not necessary to guide the sightline since it is sufficient to detect a reaction delay. However, the validity must be determined by making the subject respond to the target to prevent a false response. In this article, the measurement device’s center is called the gazing point instead of the fixation point. It is not necessary to fix the sightline by fixating on the center of the measurement device; rather, the sightline is guided to the center of the measurement device in response to the target.

The eye-guided scotoma detection method is a technique to measure the entire visual field while moving the sightline. As shown in Fig 2, the target is displayed alternately at the gazing point and the measuring point in the following steps:

**Fig 2.**
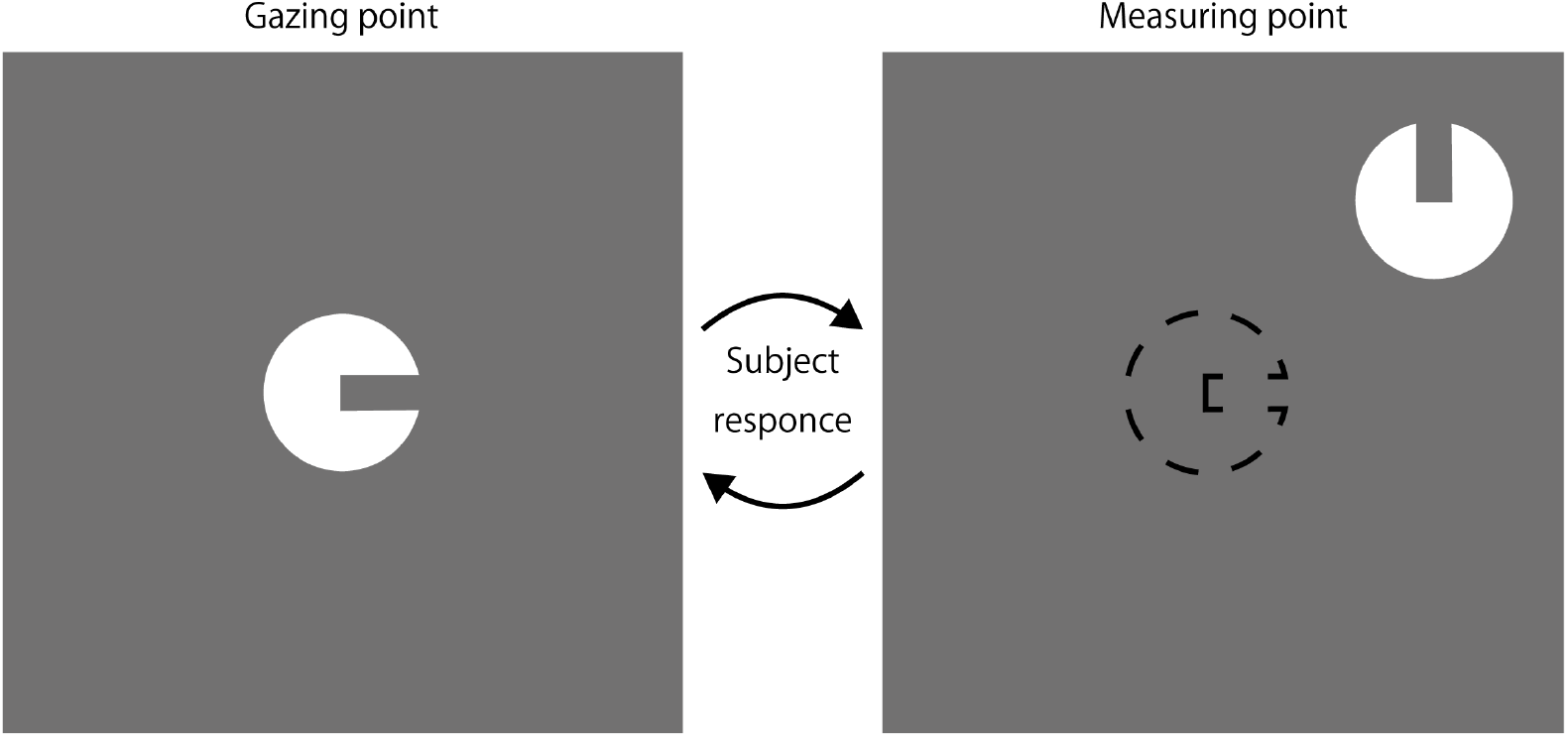
Steps of measurement by the eye-guided scotoma detection method. Guide the sightline by displaying the targets alternately at the gazing point and the measuring point and making the subject respond to the target.

1. Display the target at the gazing point, and proceed to 2 when the subject responds by matching the sightline to the target.
2. Display the target at the measuring point, and proceed to 3 when the subject responds by matching the sightline to the target or when the time expires.
3. Proceed to 1 if measurements are not completed for all the measuring points. Otherwise, terminate the measurement.

The display of the measuring point and the gazing point changes according to the subject’s response. Whether or not the subject sees the target is determined by the response time between the response at the gazing point and the response at the measuring point. Unlike the conventional perimetry, the eye-guided scotoma detection method does not require the subject to fixate at the center of the visual field and thus should reduce the burden on the subject.

### Setting the target and measuring point coordinates

Due to the eye’s spatial summation, the Landolt ring [12] has fewer stimuli to the eyes than the circle optotype since the Landolt ring is smaller stimuli areas. Therefore, the target used in this study to prevent stimuli areas loss is a clipped circle optotype (CCO), as shown in Fig 3. The size of the CCO is converted based on the target area *S*(*mm*^2^) used with HFA so that the viewing angle is the same. Convert the radius *r*(*mm*) of the CCO supposing that the viewing distance with HFA is 300(*mm*), and the viewing distance with the eye-guided scotoma detection method is *d*(*mm*) according to Eq (1).

**Fig 3.**
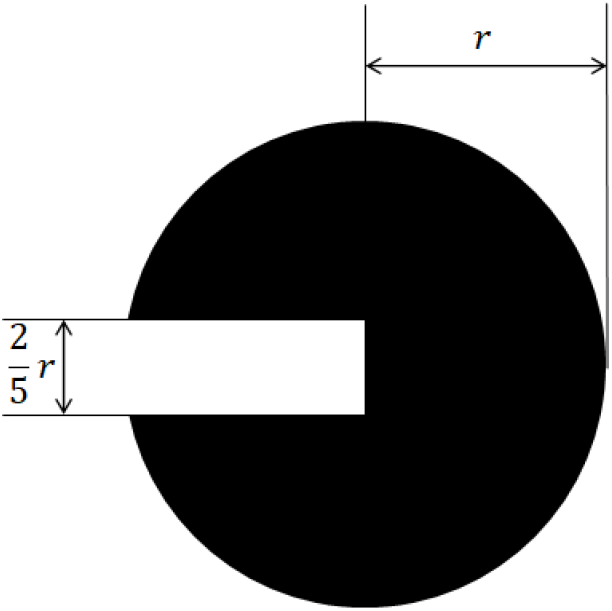
Clipped circle optotype (CCO). A circle with a diameter of 2*r* and a notch with a width of 2*r/*5 and a depth of *r*.

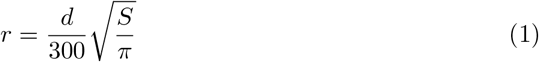

The clipping width of the CCO is 2*r/*5, and the depth is *r* as with the Landolt ring [12]. Since the CCO has a larger drawn area than the Landolt ring, the changes in measurement sensitivity due to spatial summation from the circular targets used with HFA should be smaller.

The measuring point coordinates used in this article consist of those of the center 24-2 program used with HFA [13] and the coordinates on which the measuring points are placed at the scotomas. Fig 4 shows the measuring point coordinates. The triangular indices are the measuring points corresponding to the Mariotte scotomas, and they are placed at intervals of 1 degree. These indices are the measuring points under the assumption that the center of the coordinate in the figure is the gazing point.

**Fig 4.**
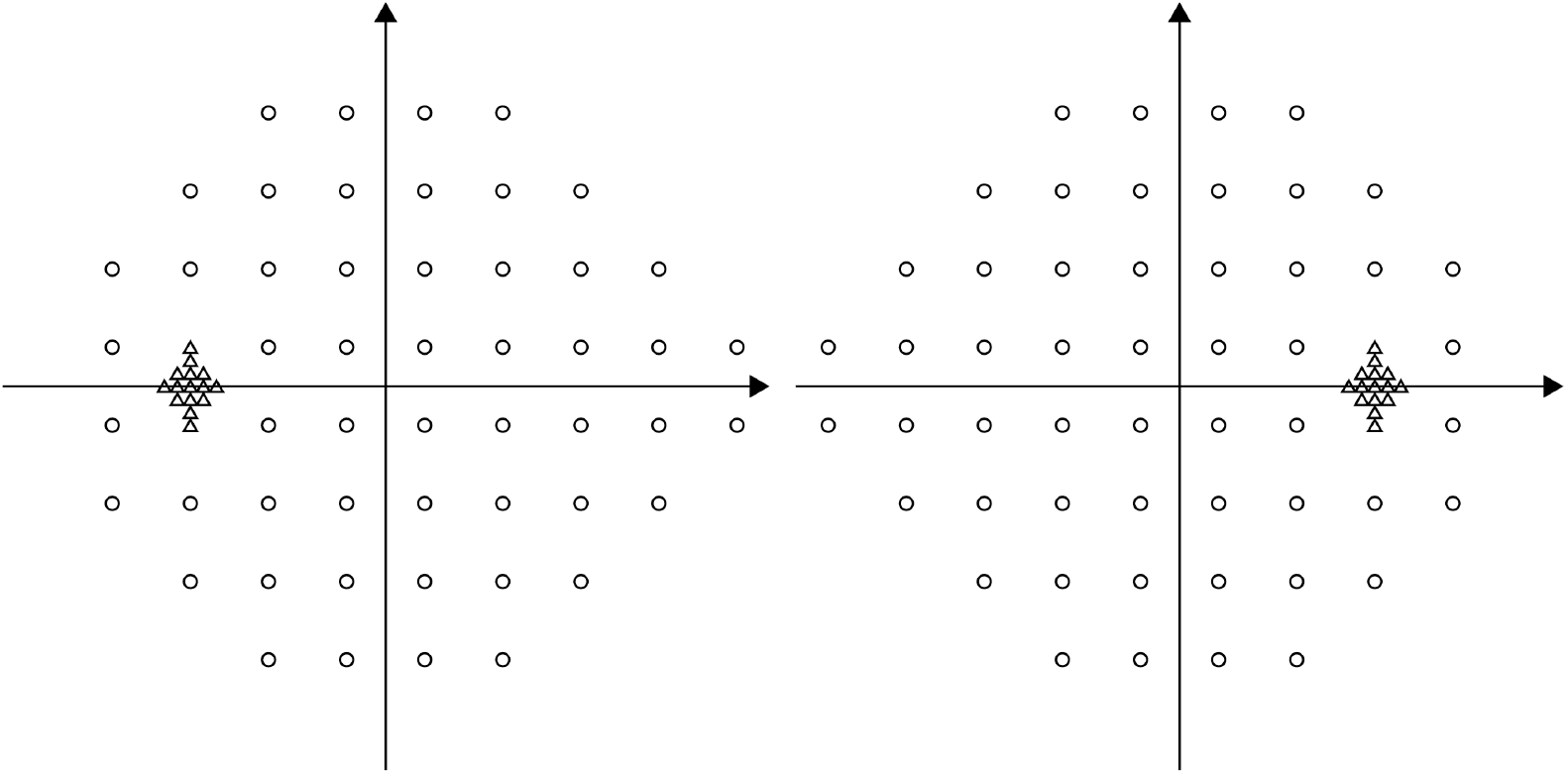
Measuring point coordinates for both eyes. In addition to the measuring point coordinates of the center 24-2 program used with HFA, the coordinates of numerous other measuring points that are placed to detect the scotomas are also used. The measuring point coordinates on the left are for the left eye, and the coordinates on the right are for the right eye.

### Measurement settings

With the eye-guided scotoma detection method, the subject enters the target’s direction displayed on the screen using an input device, such as a joystick. When the target is displayed at the gazing point, the response is considered to be correct if the direction of the notch on the CCO matches the inputted direction. When the target is displayed at the measuring point, the response is considered correct even if the direction of the notch on the CCO is different from the inputted direction. If the subject does not respond within a certain time at the measuring point, the measurement proceeds to the next gazing point as the time expires. With the eye-guided scotoma detection method, the response time is the time between the subject’s response at the gazing point and the response at the measuring point. By analyzing the response time, we determine whether sensitivity can be perceived at the measuring point. With the eye-guided scotoma detection method, to determine whether or not the subject can perceive the target, we assume a probability-of-seeing curve, as shown in Fig 5, like that used with HFA. Therefore, the probability of correct response to the measurement sensitivity at each measuring point is assumed to be *p* ≥ 50%.

**Fig 5.**
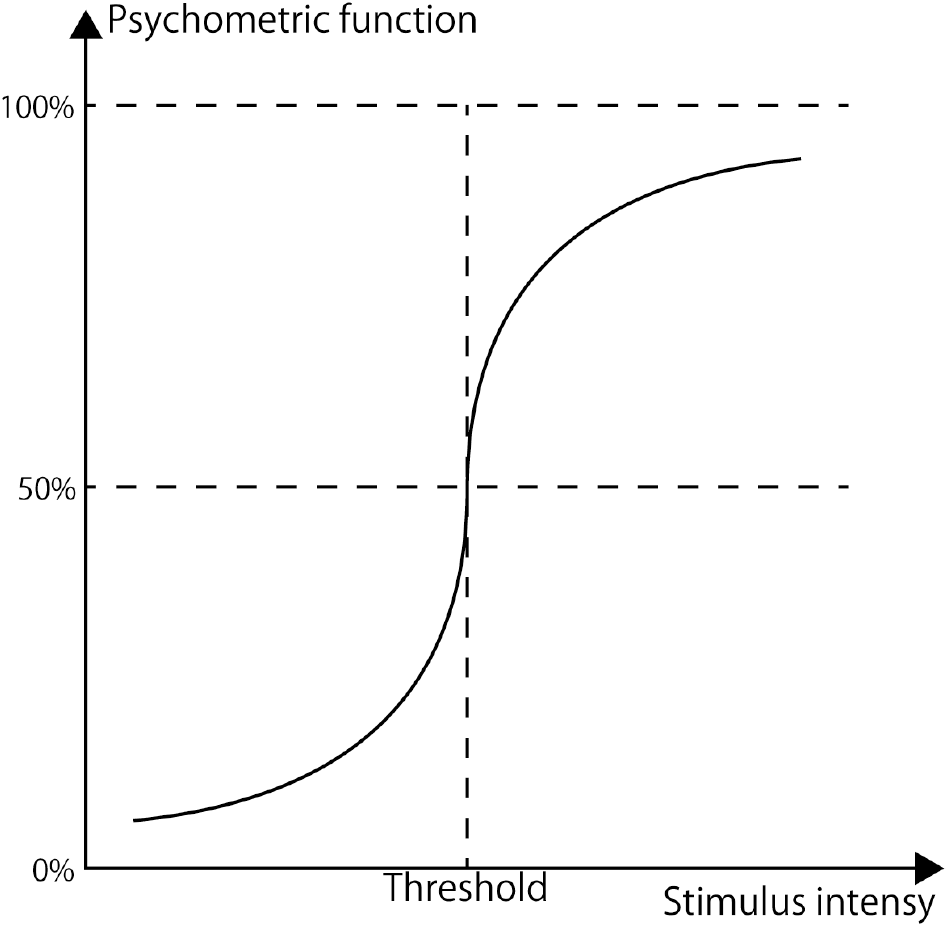
Probability-of-seeing curve. The probability-of-seeing curve assumes that the probability of perceiving the stimulus intensity corresponding to the true retinal sensitivity is 50%. The probability of perception increases as the stimulus intensity increases. The response time is shorter when the target is perceived correctly, and the response time is longer when the target is not perceived.

In this article, we propose a technique for measurement using a single sensitivity and determine whether or not the sensitivity is visible. At each measuring point, measurement is performed three times to determine whether or not the measurement sensitivity is visible based on the majority rule.

### Scotoma detection

In this study, scotomas are detected by the following two steps. In step 1, classify the response by each measurement into a scotoma and non-scotoma based on the eccentricity and response time. In step 2, evaluate the data of three measurements at each measuring point based on the majority rule according to the classification in step 1. The eccentricity is the angle of eye movement when it moves on a straight line to the coordinates of (*θ*_*x*_, *θ*_*y*_) when the gazing point is at an angle of (0, 0). The eccentricity *θ* is derived by Eq (2).

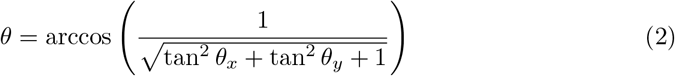

Fig 6 shows the chart of the eccentricity and response time for the data measured with the eye-guided scotoma detection method. In Fig 6, the horizontal axis is the eccentricity, and the vertical axis represents the response time. The triangular indicators are the measurement data for the Mariotte scotomas, and the circular indicators represent the other measurement data.

**Fig 6.**
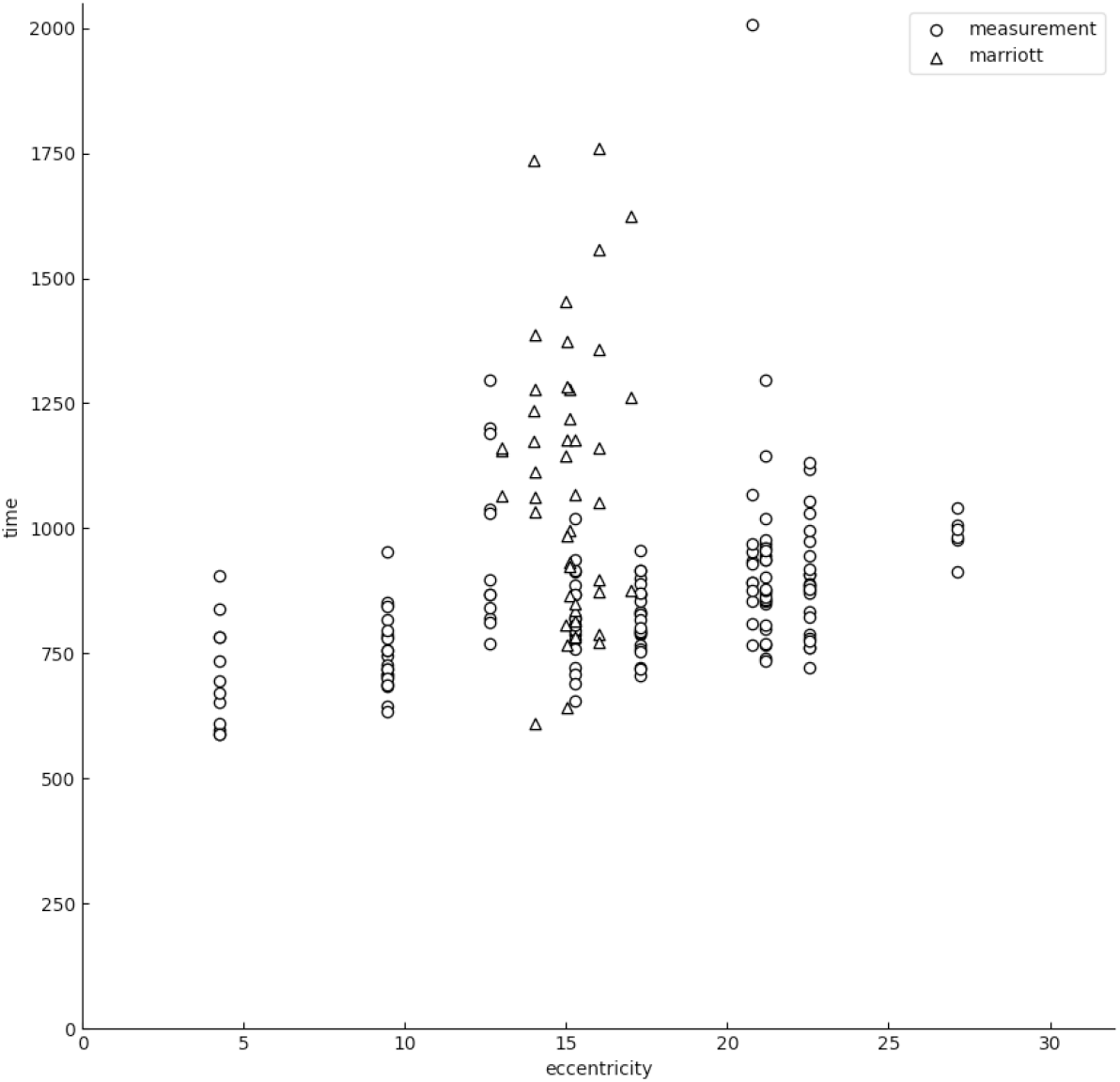
Eccentricity and response time. ⃝ represents a measuring point and △ represents the response time at the Mariotte scotoma. The Mariotte scotomas, scotomas, are plotted prominently on the upper side of the chart as delay in response occurs. ⃝ located at 2000 msec is presumed to be an outlier and not accurately measured.

With the eye-guided scotoma detection method, the response time *T* is assumed to be determined by Eq (3).

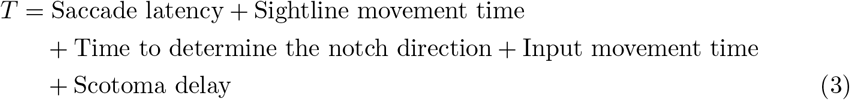

The Saccade latency, the time to determine the notch direction, and the input movement time are assumed to follow the normal distribution. The sightline movement time is assumed to depend on the eccentricity because it is the time required for eye movement. On the other hand, the scotoma delay is assumed to be close to 0 when the subject can perceive the target, and large when the target is not perceived. In this study, we propose two separation models separating the data of each measurement based on the model of Eq (3). With either model, the line of separation is finally determined, and the line of separation is used as the scotoma separation line. The measuring point at which *T* is smaller than the determined scotoma separation line is judged to be a scotoma, and the measuring point at which *T* is larger than the scotoma separation line is judged to be a non-scotoma.

### Scotoma classification by regression line

First, exclude timed-out measurement data to remove outliers from the response time. In this study, we exclude measurement data that response time upper than 2,000 msec as timed-out. Then, calculate the median value *t*_*med*_ with the eccentricity within 5 degrees and the maximum response time *t*_*max*_ at a Mariotte scotoma, and extract the data with the response time *t* within the following range.

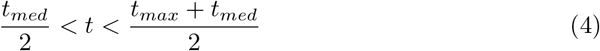

Calculate the regression line for the extracted data. The slope of this regression line is assumed to be *a* and the intercept to be *b*. Assuming that the slope corresponds to the sightline movement time in Eq (3), subtract the slope and the sightline movement time at the eccentricity *θ* from the response time at each measurement, and calculate the standard deviation *s* for the values. The lines intercepted with the standard deviation and one-sided confidence intervals of 90%, 95%, 97.5%, and 99% are used as the scotoma separation lines. The one-sided confidence intervals of 90%, 95%, 97.5% and 99% have confidence coefficients of 1.28, 1.65, 1.96 and 2.33, respectively. With the confidence coefficient *r*, the scotoma separation line, *l*_*s*_ is Eq (5).

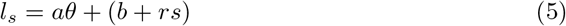

### Scotoma classification by clustering and regression line

To remove outliers as with the regression line, exclude timed-out data and extract the data that meet the conditions in Eq (4) and the data at Mariotte scotomas. The response time of the extracted data is clustered into two groups using unsupervised variational inference with the Gaussian Mixture Model (GMM) [14]. Then, calculate the regression line for the group with the faster mean response time among the clustered groups, and calculate the scotoma separation line by Eq (5). Fig 7 shows the results obtained by applying this technique to Fig 6.

**Fig 7.**
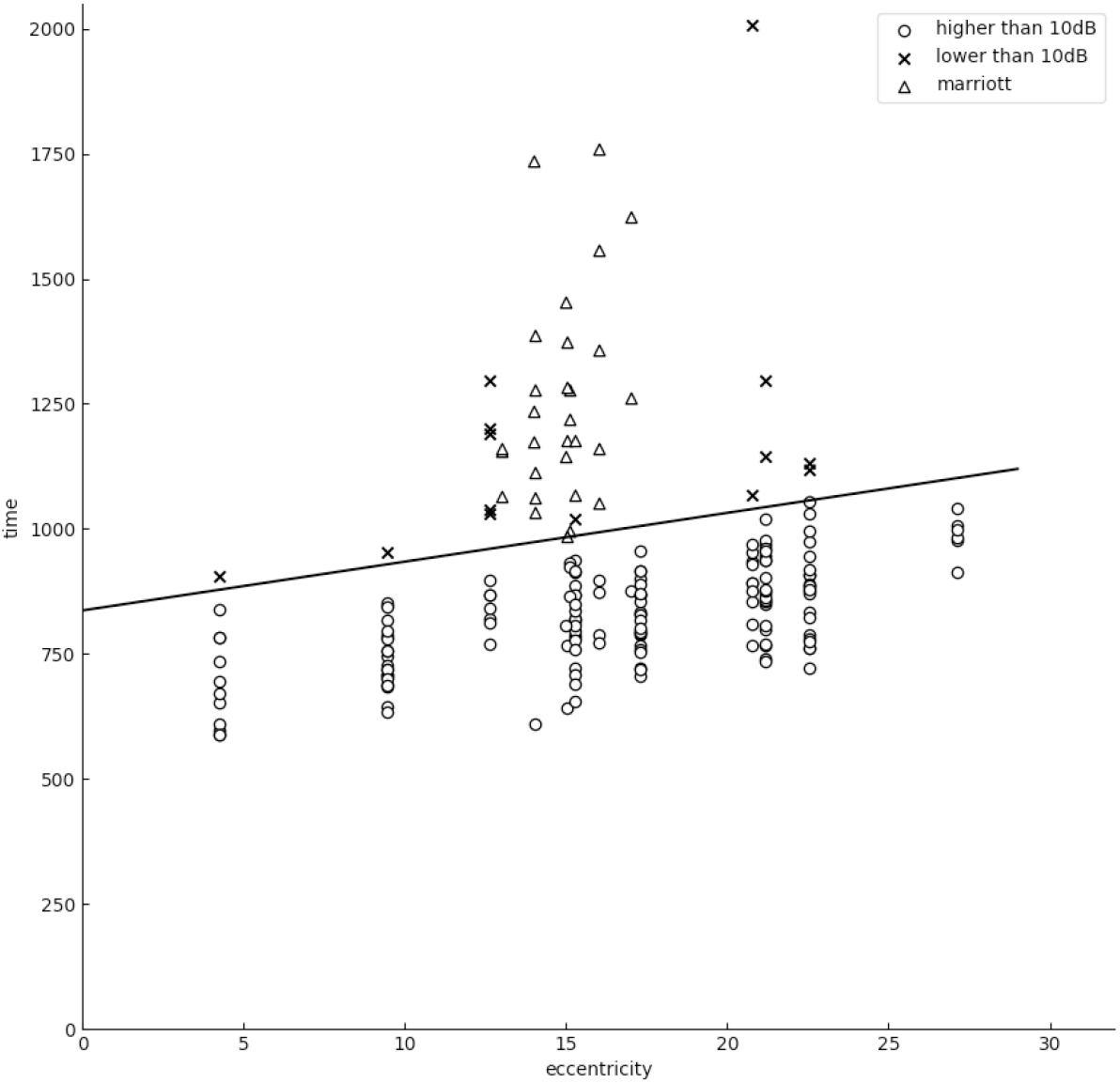
Separation by scotoma separation line. This figure shows how the scotomas and non-scotomas are determined by the scotoma separation line. The measuring points in the area above the scotoma separation line are determined to be scotomas and marked with ×. The measuring points in the area below the scotoma separation line are determined to be non-scotomas and marked with ⃝ △.represents a Mariotte scotoma.

## Evaluation

Experiments that evaluate the proposed technique were performed with the approval of the Ethics Review Board (UMIN ID: UMIN000038206) at Findex Inc.

### Subjects

The experiments were performed on 80 eyes in a total of 40 subjects for volunteers consisting of 28 males and 12 females aged 25 to 77 years (mean age: 44.2 years old; standard deviation: 14.8 years). Subjects were fully informed about this study and asked to give their voluntary consent. Table 1 shows the inclusion criteria and exclusion criteria for the experiments.

**Table 1.** Inclusion and exclusion criteria for experiments.

|  |  |
| --- | --- |
| **Inclusion criteria** | · Age of 18 years or older<br>· Patients who can provide informed consent to participate |
| **Exclusion criteria** | · Patients who reported previously having abnormal findings related to visual fixation |

These inclusion criteria and exclusion criteria were checked before the experiments.

Measurements were not performed on subjects who were totally blind or who had a disease that could affect their ability to gaze at the target.

### Experiment protocol

In this experiment, measurements were performed using an HFA II 740i and an eye-guided scotoma detection method in a dark room. Measurements were performed for visual acuity and corrected visual acuity with HFA followed by eye-guided perimetry. HFA was performed using an attached trial frame, while the eye-guided scotoma detection method used an eyeglass-formed trial frame to correct the visual acuity. In the diagnosis of glaucoma, it is considered standard practice with HFA to use a target with a viewing angle of 0.431 degrees,, which corresponds to the Goldmann perimeter’s target size III [15]. For HFA we used a target of size III to measure the center 24-2 program using SITA-Standard or SITA-Fast as the measurement algorithm.

As an eye-guided scotoma detection method, we set up a display, joystick, and chin support on the table, as shown in Fig 8. The distance from the display to the chin support was adjusted to 400 mm. The subject sat in a chair with the chin on the chin support and the head fixed to face the display. The subject covered one eye with an eye patch to shield it. The display used for the measurements was a SONY KJ-49X9500G with the following specifications: pixel count: 3840 pixels (H) × 2160 pixels (V); pixel pitch: 0.028 mm (H) × 0.028 mm (V); display surface: 107.4 mm (H) × 60.4 mm (V); display colors: 16.77 million colors; viewing angle (vertical): 178 degrees; viewing angle (horizontal): 178 degrees; maximum brightness: 700 cd/*m*^2^; and response rate: 6 ms. The personal computer used was an HP ZBook 15 G5 (CPU: Intel Core i7 8750H; memory: 16GB; HDD: 1TB; OS: Windows 10 Pro) running a measurement program developed by Microsoft Visual Studio 2015 C#. The data input device consisted of an Arduino Uno R3 joystick lever from Sanwa Denshi, a JLF-H joystick harness from Sanwa Denshi, and a YM-300 YM thin case from TAKACHI.

**Fig 8.**
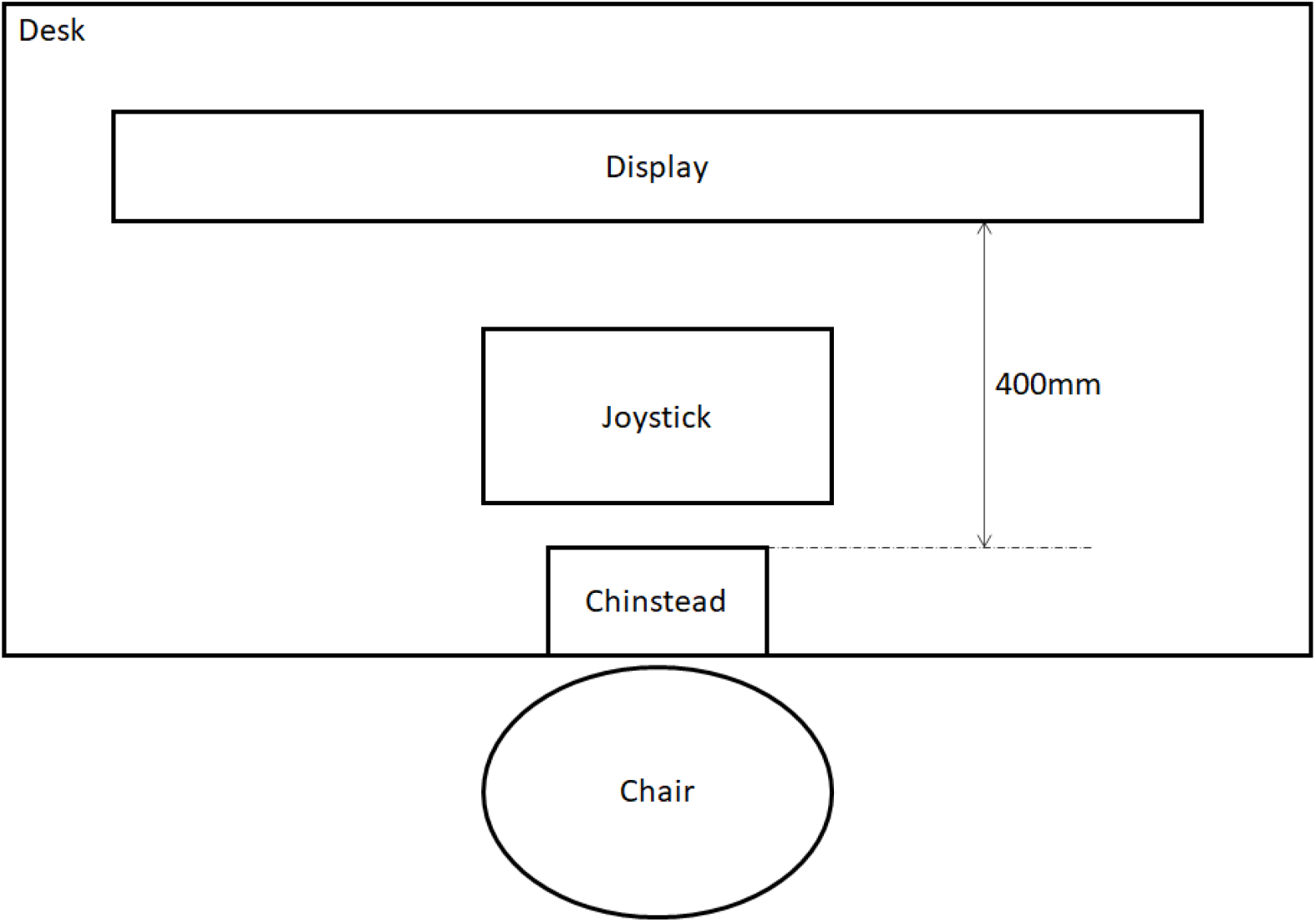
Constructed measurement system (eye-guided scotoma detection method). A display, joystick, and chin support are set up on a table, and the display and chin support are placed at a distance of 400 mm.

With the eye-guided scotoma detection method, measurement was performed at a single sensitivity of 10 dB on the measuring point coordinates in Fig 4. The target brightness was 1026.0 asb, which is equivalent to 10 dB, and the background brightness was 30.6 asb, which is the same as that of the HFA background. We compared HFA and the eye-guided scotoma detection method at each measuring point.

### Data Analysis

Evaluations were performed using the sensitivity, accuracy, false-positive rate, and false-negative rate, and the detection rate of Mariotte scotomas. In this article, a sensitivity of less than 10 dB with HFA was considered positive; a sensitivity of 10 dB or more with HFA was considered negative. In addition, the measuring point was considered to be positive if it was determined to be a scotoma by the eye-guided scotoma detection method, and it was considered negative when the measuring point was determined to be a non-scotoma. True-positive (TP), false-positive (FP), false-negative (FN), and true-negative (TN) were defined as follows:

#### True-positive *TP*

The number of measuring points that are positive with HFA and positive with the eye-guided scotoma detection method.

#### False-positive *FP*

The number of measuring points that are negative with HFA and positive with the eye-guided scotoma detection method.

#### False-negative *FN*

The number of measuring points that are positive with HFA and negative with the eye-guided scotoma detection method.

#### True-negative *TN*

The number of measuring points that are negative with HFA and negative with the eye-guided scotoma detection method.

In this case, the *Sensitivity, Accuracy*, false-positive rate (*FPr*), and false-negative rate (*FNr*) are calculated as follows;

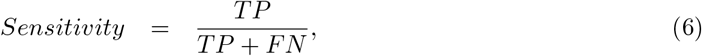

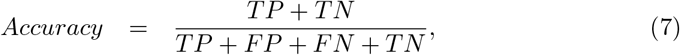

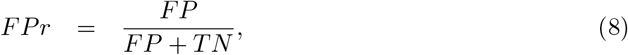

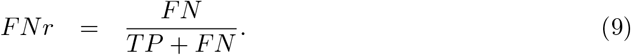

For the sensitivity, accuracy, false-positive rate, and false-negative rate, we compared the results at 52 measuring points, excluding Mariotte scotomas, with the results by HFA. In this experiment, we calculated the detection rate *M* @1 under the assumption that the Mariotte scotoma was detected when the result was determined to be positive at at least one of the measuring points corresponding to Mariotte scotomas in Fig 4. Also, we calculated the detection rate *M* @5 for Mariotte scotomas when the result was determined to be positive at 5 or more points.

## Results

Of the 80 eyes of 40 subjects included in the measurement, 2 eyes were excluded from the measurement because the subjects had a disease that affected their gazing at the target. Therefore, 78 eyes of 40 subjects were included in the analysis. The mean MD of HFA results was −1.28 dB, the standard deviation was 3.40, and the minimum was 417.99. As a result, 11 eyes had a measuring point determined to be positive, and, of these 11 eyes, 6 eyes of 5 subjects had at least 3 positive points. The 6 eyes with at least 3 positive points were considered low-sensitivity eyes, and the other eyes were handled as sighted eyes. The mean MD of low-sensitivity eyes was -10.70 dB (standard deviation was 5.72 dB), and the mean MD of sighted eyes was −0.60 dB (standard deviation was 1.68 dB). Fig 9 shows the results of HFA and the eye-guided scotoma detection method for subjects with many positive measurement points. Table 2 shows the evaluation indices for the measurement results calculated for all eyes, sighted eyes, and low-sensitivity eyes.

**Table 2.** Comparison of measurement results on a total of 78 eyes between HFA and the eye-guided scotoma detection method (%)

|  | Methods | *Sensitivity* | *Accuracy* | *FPr* | *FNr* | *M@1* | *M@5* |
| --- | --- | --- | --- | --- | --- | --- | --- |
| All results (n=78) | Regression 90% | 87.1 | 90.6 | 9.4 | 12.9 | 100.0 | 83.3 |
|  | Regression 95% | 87.1 | 92.8 | 7.2 | 12.9 | 100.0 | 82.1 |
|  | Regression 97.5% | 81.2 | 94.0 | 5.7 | 18.8 | 100.0 | 74.4 |
|  | Regression 99% | 76.5 | 95.0 | 4.6 | 23.5 | 98.7 | 61.5 |
|  | GMM regression 90% | 91.9 | 83.2 | 17.0 | 8.1 | 100.0 | 93.6 |
|  | GMM regression 95% | 90.7 | 86.9 | 13.1 | 9.3 | 100.0 | 88.5 |
|  | GMM regression 97.5% | 89.5 | 89.3 | 10.7 | 10.5 | 100.0 | 87.2 |
|  | GMM regression 99% | 87.2 | 91.3 | 8.6 | 12.8 | 100.0 | 84.6 |
| Sighted results (n=72) | Regression 90% | 87.1 | 90.6 | 9.4 | 12.9 | 100.0 | 83.3 |
|  | Regression 95% | 87.1 | 92.8 | 7.2 | 12.9 | 100.0 | 82.1 |
|  | Regression 97.5% | 81.2 | 94.0 | 5.7 | 18.8 | 100.0 | 74.4 |
|  | Regression 99% | 76.5 | 95.0 | 4.6 | 23.5 | 98.7 | 61.5 |
|  | GMM regression 90% | 91.9 | 83.2 | 17.0 | 8.1 | 100.0 | 93.6 |
|  | GMM regression 95% | 90.7 | 86.9 | 13.1 | 9.3 | 100.0 | 88.5 |
|  | GMM regression 97.5% | 89.5 | 89.3 | 10.7 | 10.5 | 100.0 | 87.2 |
|  | GMM regression 99% | 87.2 | 91.3 | 8.6 | 12.8 | 100.0 | 84.6 |
| Low-sensitivity results (n=6) | Regression 90% | 88.8 | 76.0 | 28.4 | 11.3 | 100 | 100 |
|  | Regression 95% | 88.8 | 78.5 | 25.0 | 11.3 | 100 | 83.3 |
|  | Regression 97.5% | 83.8 | 79.8 | 21.6 | 16.3 | 100 | 83.3 |
|  | Regression 99% | 78.8 | 82.1 | 16.8 | 21.3 | 100 | 50.0 |
|  | GMM regression 90% | 92.6 | 69.6 | 38.5 | 7.4 | 100 | 100 |
|  | GMM regression 95% | 91.4 | 74.0 | 32.0 | 8.6 | 100 | 83.3 |
|  | GMM regression 97.5% | 91.4 | 76.3 | 29.0 | 8.6 | 100 | 83.3 |
|  | GMM regression 99% | 88.9 | 76.3 | 28.1 | 11.1 | 100 | 83.3 |

*Sensitivity* is the sensitivity, *Accuracy* is the accuracy, *FPr* is the false-positive rate, *FNr* is the false-negative rate, *M@1* is the rate of detecting at least one Mariotte scotoma, and *M@5* is the rate of finding 5 or more positive measuring points corresponding to Mariotte scotomas. Regression represents the calculation result of the separation line with each confidence interval for the analyzed results using regression analysis alone. GMM regression means the calculation result of the separation line with each confidence interval for the analyzed results using GMM and regression analysis.

**Fig 9.**
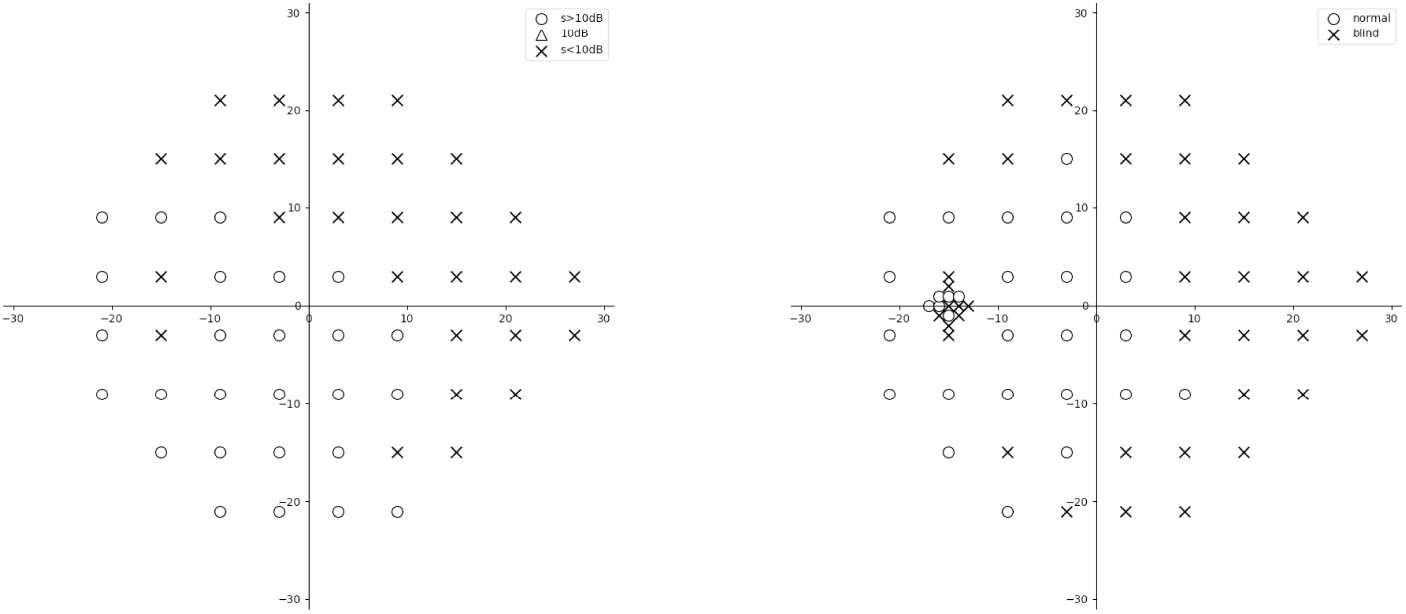
Measurement results with HFA and the eye-guided scotoma detection method. The left figure shows the measurement results with HFA, and the right figure shows the measurement results with the eye-guided scotoma detection method. × represents a measuring point at which the sensitivity was less than 10 dB and determined to be positive. ⃝ represents a measuring point at which the sensitivity was 10 dB or higher and determined to be negative.

Since the proposed eye-guided scotoma detection method is a measurement method with a single sensitivity for screening, it is necessary to detect visual field defects with high accuracy. Therefore, this method appears to be appropriate as a technique with high sensitivity (*Sensitivity*), a low false-negative rate (*FNr*), and high accuracy. As shown in Table 2, it can detect Mariotte scotomas, since the *M* @1 is 100% except for 99% for regression; therefore, it should be capable of detecting scotomas with high accuracy depending on how the scotoma separation line is determined. These findings suggest that it is the best practice to use the combination of GMM and regression lines with a 90% confidence interval by setting a scotoma separation line with high sensitivity (*Sensitivity*), a low false-negative rate (*FNr*), and high *M* @1 and *M* @5g rates.

## Discussion

We developed an eye-guided scotoma detection method and applied it to check for the presence of scotoma in 78 eyes of 40 subjects. With the scotoma detection using the scotoma separation line depending on eccentricity and response time, we were able to control the false-negative rate to less than 10% while maintaining the Mariotte scotoma detection rate of 100%. Using our eye-guided scotoma detection method, the scotoma classification was more accurate with a lower false-negative rate at any confidence interval compared with the classification with the regression line alone.

Since the eye-guided scotoma detection method is meant for mass screening, it is important to have a low false-negative rate to avoid missing any scotomas. Also, the high detection rate of Mariotte scotomas suggests the high reliability of scotoma detection in the proposed method. The combination of high accuracy and a high detection rate of Mariotte scotomas also indicates high validity. In this experiment, scotomas were detected in subjects with sighted eyes, but if no scotomas were detected with HFA, the accuracy could not be determined as calculated by Eq (6). In that case, the performance could be evaluated by determining whether the false-positive rate becomes lower when it is calculated by Eq (8).

The differences by confidence interval in the scotoma classification by the combination of GMM and regression line show that the results with a confidence interval of 90% are most accurate with the lowest false-negative rate. By calculating the regression line after clustering by GMM, GMM excludes the data that cannot be excluded by Eq (4). Therefore, the combination of GMM and regression line might be more accurate than a regression line alone. A comparison of the positive measuring points with HFA and the eye-guided scotoma detection method shown in Fig 9 shows that the positive points are similar in terms of shape and location, suggesting this classification method’s validity. Therefore, the eye-guided scotoma detection method shows a high concordance in the classification results with HFA at a boundary of 10 dB, suggesting that it is possible to perform screening with a single sensitivity. It is also suggested that the sensitivity can be measured by the eye-guided scotoma detection method, because the results were consistent when divided by sensitivity in this experiment.

Furthermore, the eye-guided scotoma detection method can also be used to evaluate the reliability of measurement using the measurement of Mariotte scotoma. The measurement with HFA uses the fixation loss, false-positive rate, and false-negative rate of the test as reliability indices. The fixation loss evaluates eye movements, which shift the position of gaze away from the fixation point. Hence, fixation loss is important for the reliability of the visual field test [16]. On the other hand, the eye-guided scotoma detection method needs to guide the eyes to the position of the gazing point and measuring point rather than to a fixation point. The eye-guided scotoma detection method evaluates the guide loss using the measurement of Mariotte scotomas. If the patient responds to the stimulus that presented the Mariotte scotoma, the eye-guided scotoma detection method is not sufficient to guide the eyes to the gazing point and measuring point. Moreover, if the eye-guided scotoma detection method can detect Mariotte scotomas, the proposed method may detect scotomas. Therefore, the eye-guided scotoma detection may be appreciated as a reliable scotoma detection method if it can detect Mariotte scotomas.

The Goldmann perimeter is unlikely to be available since it is a one-on-one examination, and the examiner requires about 30 minutes of examination time per eye [17]. HFA is unlikely to be available due to its lack of portability: it is impractical to provide the necessary number of units for mass screening due to high costs [18]. The Amsler chart is convenient for scotoma detection, but it is unlikely to be available since it is subjective and requires subjects to correctly understand the nature of the test [19]. The blackboard perimetry and confrontation test are unlikely to be available since they need to be performed one-on-one by an examiner who has knowledge and skills in visual function assessment. The common visual field examination methods require the subject to stare at the center point of the perimeter without moving his/her sightline and respond to a glowing point around the center, for example, by pressing a button. It is very stressful for the subject to fixate on a center point, as he/she is forced to do during perimetry, and to be prohibited from moving the eyes to see something glowing; it is human nature to move the eyes to see whether there really is something glowing. In addition, perimetry requires a large and complicated system consisting of dedicated devices to project the target and monitor fixation and a dark room. The measurement itself requires a specialized medical professional to operate the devices and *thus* imposes a heavy burden as he/she must monitor the subject’s fixation constantly. In contrast, with the eye-guided scotoma detection method, the subject is inevitably guided to fixate at the center of the screen (gazing point) in order to distinguish the direction of the notch on the CCO; therefore, this method requires neither monitoring of fixation by the examiner nor a computerized fixation monitoring function. The measurement system (perimeter) cost should be reduced since fixation monitoring is not required, suggesting that a single examiner can perform the examination.

The eye-guided scotoma detection method has excellent portability because it can be configured using commonly used information devices. In particular, the system can be rendered more portable by replacing the display with a head-mounted display that requires no large display or chin support. A low-cost measurement system can be created with common information devices. Also, because the examination procedure is simple and easy to understand, the examiner does not have to assist subjects on a one-to-one basis, and the system is powerful enough to detect Mariotte scotomas and scotomas, it could be used as a method of mass screening at schools and workplaces by providing the necessary number of measurement units. Finally, we should note that there are important two limitations to the eye-guided scotoma detection method. The eye-guided scotoma detection method can ‘t measure the eye, which has abnormal findings related to visual fixation, such as central scotoma, due to not judge the optotype direction. Besides, in this study, insufficient verification of various abnormal visual field cases, and we need to verify the eye-guided scotoma detection method with the various subjects.

## Conclusion

For the eyes determined to be low-sensitivity eyes, a similar distribution of positive points was observed with HFA and the eye-guided scotoma detection method. The eye-guided scotoma detection method has the following advantageous characteristics: 1) It enables the configuration of a transportable measurement system; 2) It enables the creation of a low-cost measurement system; 3) It enables the performance of examinations by a single examiner. These findings suggest that the system could be used as a method of mass screening for glaucoma at schools and workplaces by providing the necessary number of measurement units.

## Data Availability

The data gathered during and analyzed in the current study are not publicly available to the restrictions of the commercially but are available from the corresponding author on reasonable request.

https://www.kaggle.com/shozosaeki/visual-field-testing-experiment

## Acknowledgments

This work was supported by JSPS KAKENHI Grant Number JP18K11528.

## Author Contributions

**Conceptualization:** Nobuyuki Takahashi, Shozo Saeki, Minoru Kawahara, Hirohisa Aman

**Data curation:** Nobuyuki Takahashi, Shozo Saeki

**Formal analysis:** Shozo Saeki

**Funding acquisition:** Minoru Kawahara

**Investigation:** Nobuyuki Takahashi, Shozo Saeki, Minoru Kawahara

**Methodology:** Nobuyuki Takahashi, Shozo Saeki, Minoru Kawahara, Hirohisa Aman, Eri Nakano, Yuki Mori, Masahiro Miyake

**Project administration:** Shozo Saeki, Minoru Kawahara, Masahiro Miyake

**Resources:** Shozo Saeki

**Software:** Nobuyuki Takahashi, Shozo Saeki, Hirohisa Aman

**Supervision:** Minoru Kawahara, Hirohisa Aman, Hiroshi Tamura, Akitaka Tsujikawa

**Visualization:** Nobuyuki Takahashi, Shozo Saeki, Minoru Kawahara, Hirohisa Aman

**Writing - original draft:** Nobuyuki Takahashi, Shozo Saeki, Minoru Kawahara, Hirohisa Aman

**Writing – review & editing:** Shozo Saeki, Minoru Kawahara, Eri Nakano, Yuki Mori, Masahiro Miyake, Hiroshi Tamura, Akitaka Tsujikawa

## Notes

### Competing Interest Statement

This research unrestricted research grants from FINDEX Inc.
This research relates the patent application which is "VISUAL FIELD MEASURING METHOD, VISUAL FIELD MEASURING APPARATUS, AND OPTOTYPE," US 10,687,700 B2.
Shozo Saeki has employed by FINDEX Inc.
Dr. Kawahara reports grants from JSPS KAKENHI JP18K11528 and grants from
Findex.
Dr. Miyake reports personal fees from Santen Pharm., personal fees from Nevaker, personal fees from Novartis Pharm., personal fees from Japan Alcon, personal fees from HOYA, personal fees from Kowa Pharm., personal fees from Senju Pharm., personal fees from Ellex, personal fees from Johnson & Johnson, personal fees from AMO Japan, outside the submitted work;.
Dr. Tamura reports grants from Ministry of Health, Labor, and Welfare, personal fees from Novartis, personal fees from Santen Pharmaceutical, personal fees from Bayer Yakuhin, personal fees from Suntory, outside the submitted work.
Dr. Tsujikawa reports grants and personal fees from Canon, grants from Findex, grants and personal fees from Santen Pharmaceutical, grants from Kowa Pharmaceutical, grants from Pfizer, grants and personal fees from AMO Japan, grants and personal fees from Senju Pharmaceutical, grants and personal fees from Wakamoto Pharmaceutical, grants and personal fees from Alcon Japan, grants and personal fees from Alcon Pharma, grants and personal fees from Otsuka Pharmaceutical, grants from Tomey Corporation, grants from Taiho Pharma, personal fees from Hoya, personal fees from Bayer Yakuhin, personal fees from Novartis Pharma, personal fees from Chugai Pharmaceutical, personal fees from Astellas, personal fees from Eisai, personal fees from Daiichi-Sankyo, personal fees from Janssen Pharmaceutical, personal fees from Kyoto Drug Discovery & Development, personal fees from Allergan Japan, personal fees from Sanwa Kagaku Kenkyusho, personal fees from Nitten Pharmaceutical, personal fees from Otsuka Pharmaceutical, outside the submitted work.

### Clinical Trial

Trial ID is UMIN000038206

### Funding Statement

Minoru Kawahara.: JSPS KAKENHI Grant Number JP18K11528,
https://www.jsps.go.jp.
Minoru Kawahara, Hiroshi Tamura, and Akitaka Tsujikawa received funding from FINDEX Inc.
(https://findex.co.jp/en/index.html)FINDEX Inc.
FINDEX Inc. had the role which data collection and the process of proofreading the manuscript in the study.

### Author Declarations

Ethics Review Board at Findex Inc.

### Summary of Updates

We update subject's informations

